# Landscape assessment to characterize baseline access and multilevel barriers to IMProve Access to CAR-T CD19 therapy (IMPACT study) across Europe

**DOI:** 10.1101/2025.11.04.25339516

**Authors:** Aleksandra Oszer, Jacques-Emmanuel Galimard, Joseph R. Wardell, Meenakshi Devidas, Arnaud Dalissier, Antonio Perez-Martinez, Nancy S. Bolous, Szymon Janczar, Jan Styczyński, Taisiya Yakimkova, Carlos Rodriguez-Galindo, Wojciech Młynarski, Asya Agulnik, Caitlyn Duffy, Krzysztof Kałwak, Kjeld Schmiegelow, St. Jude Global, the European Group for Blood and Marrow Transplantation (EBMT) PDWP and the International BFM Study Group (I-BFM)

## Abstract

**Background/Purpose:** Chimeric Antigen Receptor-T Cell Therapy (CAR-T) has revolutionized the treatment of B-cell precursor acute lymphoblastic leukemia (B–ALL), but its global availability is limited. This study assessed current access and barriers to CAR-T CD19 cell therapy for children across Europe.

**Methods:** A country questionnaire developed by the EBMT PDWP, St. Jude Children’s Research Hospital, and IBFM assessed current access to advanced therapies for B-ALL in Europe using Qualtrics software.

**Results:** Data from 35 WHO-defined European countries (26 high-income, 9 upper middle-income) revealed a median of 5 pediatric hematology-oncology (PHO) centers per country (0.55 PHO centers/1 million inhabitants, range: 0.05-1.83). Hematopoietic stem cell transplantation (HSCT) facilities were available in 89% of countries (31/35). Sixty B-ALL cases were diagnosed annually per country (4 B-ALL children/100,000 children, range: 0.4-8.4). CAR-T CD19 therapy was available in 71% of countries; however, more than 50% of countries lacked clinical trials or international collaborations for pediatric CAR-T CD19 therapy. Most countries accepted foreign patients, but referrals remained limited, with 1-2 foreign patients treated annually per country. Seventeen countries expressed interest in a referral network, but only six had established mechanisms for domestic or international referrals.

**Conclusion:** Substantial disparities exist in access to advanced therapies for pediatric B-ALL across Europe. Although CAR-T CD19 therapy is available in most countries, gaps in clinical trials, collaborations, and referral systems limit equitable access. Efforts to improve infrastructure and establish referral networks are essential to enhance care for pediatric B-ALL patients.

## Introduction

B-cell precursor acute lymphoblastic leukemia (B - ALL) is the most common hematological malignancy in children. Although contemporary therapy achieves cure rates exceeding 90%, 15–20% of patients relapse or remain refractory to frontline treatment. For these children, salvage therapy involves intensive chemotherapy and often allogeneic hematopoietic stem cell transplantation (HSCT), which carries a substantial risk of toxicity and long-term complications.^1^ Novel immunotherapies, particularly blinatumomab and Chimeric Antigen Receptor T-cell (CAR-T) CD19 therapy and blinatumomab, have transformed the management of relapsed or refractory B-ALL by harnessing the patient’s own immune system to target leukemic cells, often with reduced systemic toxicity compared with conventional chemotherapy. The landmark ELIANA trial demonstrated the efficacy of CAR-T CD19 therapy in patients with refractory disease or multiple relapses, reporting 1-year event-free and overall survival rates of 50% and 76%, respectively.^2^ Subsequently, CAR-T CD19 therapy has rapidly emerged and been approved as a critical “last-resort” treatment for patients with primary refractory disease or after multiple and/or post-transplant relapses, but is yet to be included in 1^st^ and 2^nd^ line therapy for high risk patients.^3^

However, nearly 90% of children with cancer reside in low- and middle-income countries (LMICs), where survival rate for ALL remains significantly lower, ranging from 10% to 60%.^4^ This disparity is multifactorial, but limited access to advanced therapies such as CAR-T therapy is a major contributor and may further widen the survival gap.^5^ Global access to CAR-T CD19 therapy is constrained by both cost and infrastructure. Tisagenlecleucel (Kymriah, Novartis), the first FDA-approved CAR-T CD19 product in 2017,^6^ carries a list price of $475,000, which is prohibitive for lower-resources centers. One potential solution for LMIC centers is developing academic Point of Care CAR-T CD19 manufacturing, enabling local production of CAR-T cells at substantially reduced costs. For example, a center in India produced a CAR-T CD19 therapy for approximately $50,000, bringing this life-saving treatment closer to global accessibility.^7^ But further clinical development of CAR-T therapy to its highest potential also in earlier lines of therapy will require strict product comparability across sites.

In Europe, significant disparities exist in access to CAR-T therapy, particularly between high-income countries (HICs) and LMICs. These inequities are driven by a combination of high treatment costs, limited manufacturing capacity, and variable referral networks, leaving many children with relapsed or refractory B-ALL without timely access to this life-saving therapy.

To address these gaps, we designed the IMProve Access to CAR-T cell Therapy (IMPACT) study, a Europe-wide assessment of CAR-T CD19 therapy availability. The primary objectives were to map current access to CAR-T therapy, quantify the potential population of pediatric patients eligible for CAR-T CD19 therapy, and identify multilevel barriers to implementation. By presenting a comprehensive overview of CAR-T CD19 therapy accessibility, the IMPACT study aims to guide strategies to promote equitable access to advanced immunotherapies for children with B-ALL across diverse healthcare systems.

## Material and Methods

### Study design

This study employed a cross-sectional design to evaluate access to CAR-T CD19 therapy for pediatric patients with B-ALL across European countries. The primary aim was to assess current availability of and identify gaps in CAR-T access, in both LMICs and HICs. The study was conducted by a multidisciplinary team comprising members of the European Group for Blood and Marrow Transplantation (EBMT) Pediatric Diseases Working Party (PDWP), St. Jude Children’s Research Hospital – St. Jude Global^8^, and the International Berlin-Frankfurt-Münster (BFM) Study Group,Committee ALL. The study was guided by an implementation framework for novel therapies to facilitate translation of advanced pediatric cancer treatments into clinical practice.^9^

### Study population and sampling

All Hematopoietic Stem Cell Transplant (HSCT) centers within Europe were invited to participate. In countries lacking HSCT centers, all PHO centers were approached. Eligible participants included national coordinators for pediatric B-ALL treatment or, if no national coordinator was available, institutional PHO leads. One response per country was collected from the national coordinator where available; otherwise, one response per institution was included.

### Survey development and administration

Two complementary surveys were developed through engagement of experts from the coordinating collaborative group: an institutional assessment questionnaire (Supplement 1) and a country-level assessment questionnaire (Supplement 2). Surveys included multiple-choice questions and Likert-scale items assessing access to CAR-T therapy, clinical practice, and institutional infrastructure. Draft surveys were piloted in two HSCT and two PHO centers to ensure clarity, feasibility, and relevance; feedback was incorporated into the final versions. Surveys were developed and administered electronically via the Qualtrics platform and survey links were distributed via email through collaborative group networks. Participating centers had a 6-month period to complete the survey, with intermittent email reminders to maximize response rates. Data collection concluded at the end of October 2025.

### Data collection and variables

Survey data included:

- Access to advanced therapies and diagnostics: availability of CAR-T CD19 therapy, HSCT, blinatumomab and minimal residual disease (MRD) monitoring.
- Patient-level estimates: annual incidence of B-ALL, 5-year event-free survival, 5-year overall survival and relapse rate.
- CAR-T CD19 therapy-specific variables: type of CAR-T CD19 product administered, availability of local manufacturing facilities.
- Referral and networking pathways: existing mechanisms for referring patients to CAR-T therapy centers.
- Multilevel determinants impacting CAR-T CD19 access

Data were collected at both the institutional and national levels. In the absence of a national coordinator, institutional data were extrapolated to provide approximate country-level estimates. This approach enabled estimation of the annual number of pediatric patients in need of CAR-T therapy across participating countries. Data analysis and visualization was performed by R version 4.5.1. Descriptive statistics were used to summarize categorical data using percentages and continuous data using median (range).

### Collaboration and expert oversight

A core expert team provided methodological oversight, validated survey content, and ensured accurate interpretation of responses. Engagement with EBMT-PDWP and the International BFM Study Group facilitated connection with national coordinators and institutional leads, maximizing response rates and data completeness.

### Ethical considerations

The study was conducted in accordance with ethical principles for observational research. Approval was obtained from the Bioethics Committee at the Medical University of Lodz, Poland (RNN/56/24/KE) and from the St Jude Children’s Research Hospital, USA (IRB number: 24-1650).Participation was voluntary, and no individual patient identifiers were collected, and data were analyzed in aggregate to maintain confidentiality.

## Results

The study included participants from 35 WHO-defined European countries: 26 high-income and 9 upper-middle-income countries. National-level responses were obtained from 24 countries: Armenia, Austria, Switzerland, Czech Republic, Germany, Denmark, Spain, Estonia, Finland, France, Croatia, Italy, Kazakhstan, Lithuania, Latvia, Moldova, Netherlands, Norway, Portugal, Romania, Slovakia, Slovenia, Ukraine, and Bulgaria. Institutional-level responses were collected from 11 additional countries: Albania, Belgium, the United Kingdom, Greece, Hungary, Israel, Poland, Serbia, Turkey, Belarus, and Russia (Figure 1).

**Figure 1.**
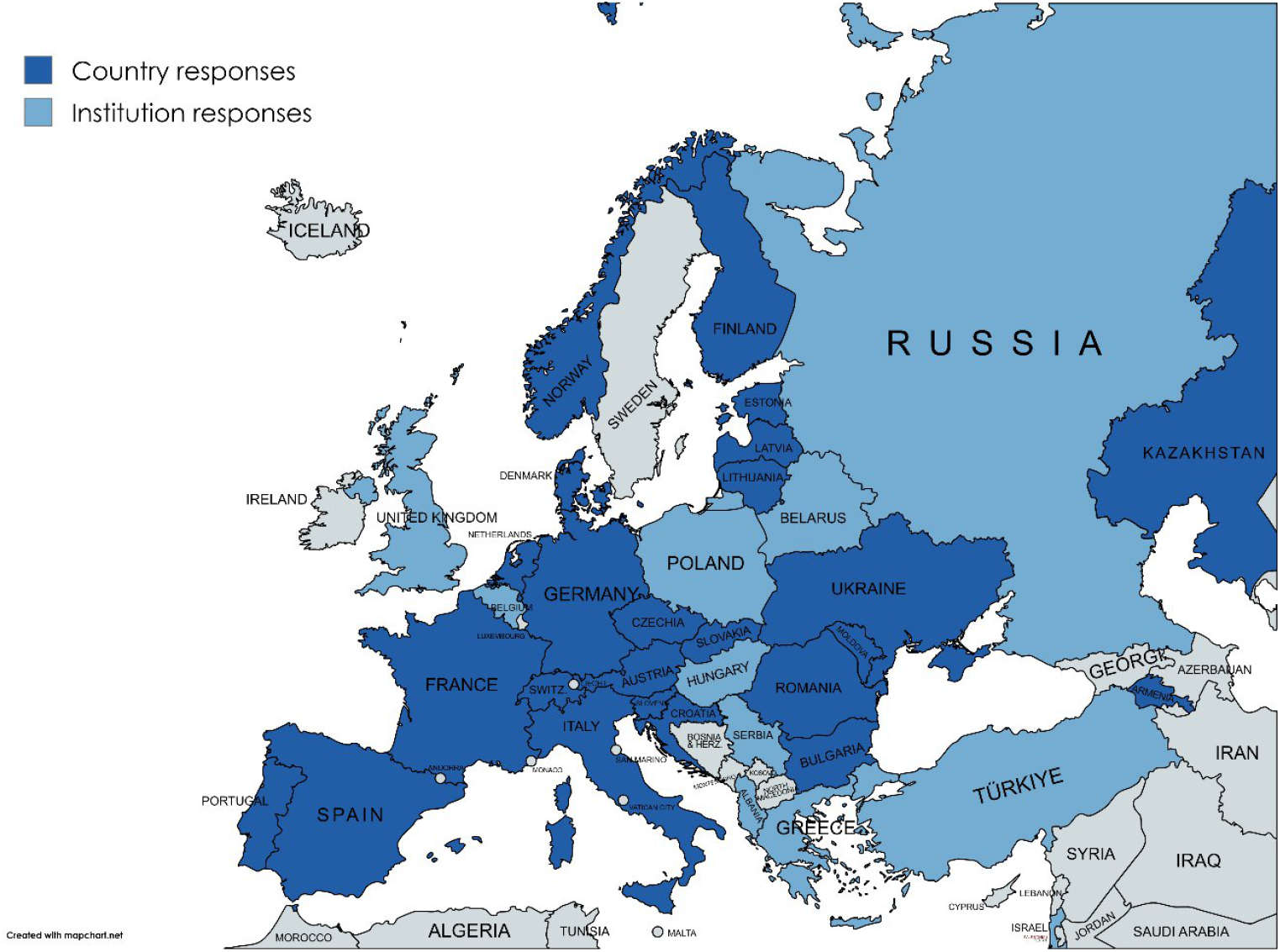
Study participation. Dark blue – National-level responses. Light blue – Institution-level responses.

Across all countries, the median number of pediatric hematology-oncology centers per country was 5, or 0.55 pediatric hematology-oncology centers/1 million inhabitants, range: 0.05-1.83 (Figure 2A). The mean annual incidence of B-ALL was 60 per country, or 4 B-ALL cases/100,000 children, range: 0.4-8.4 (Figure 2B). HICs reported a higher median incidence, 62 cases annually, compared to LMICs, 30 cases (Table 1).

**Figure 2.**
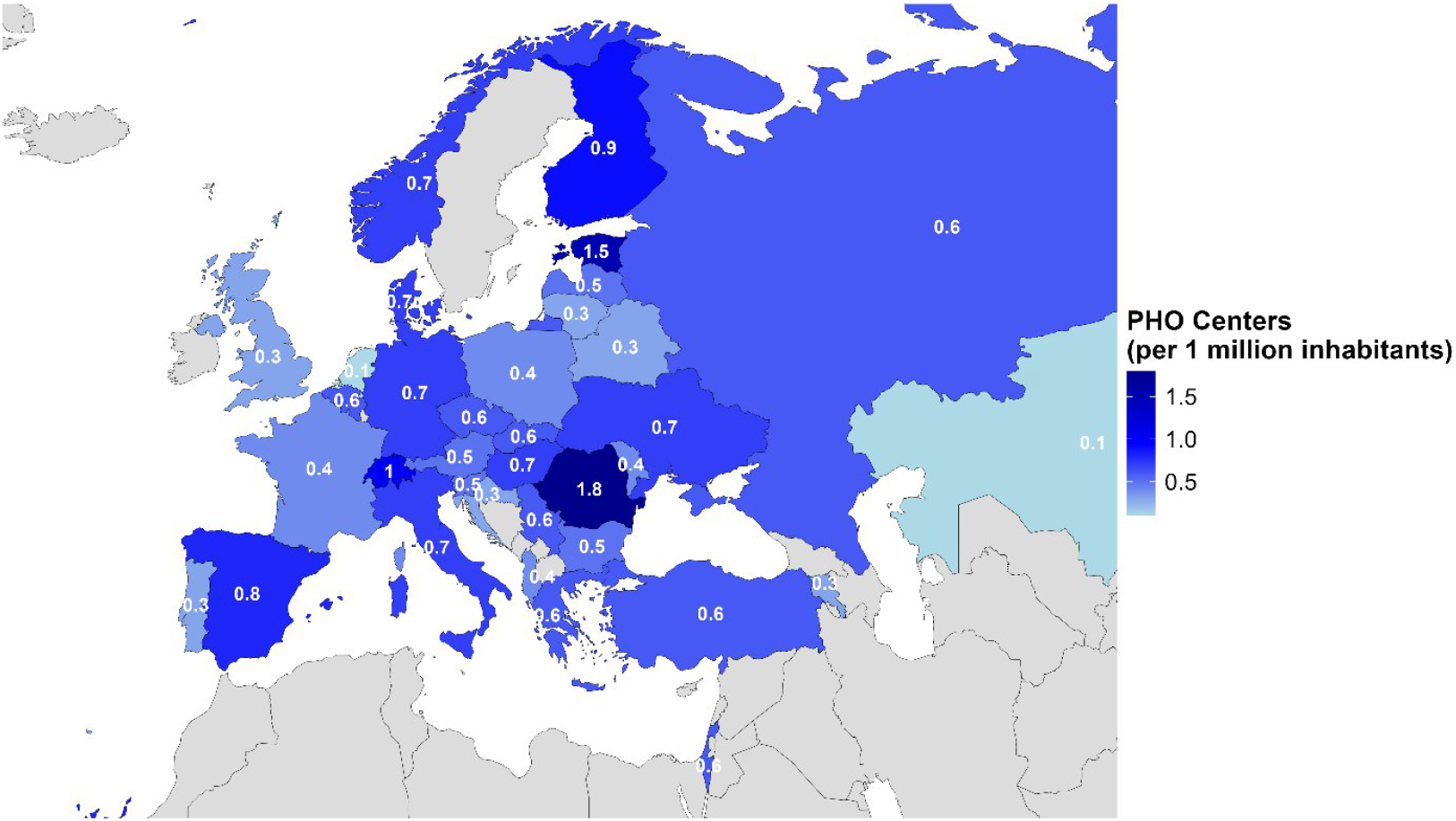
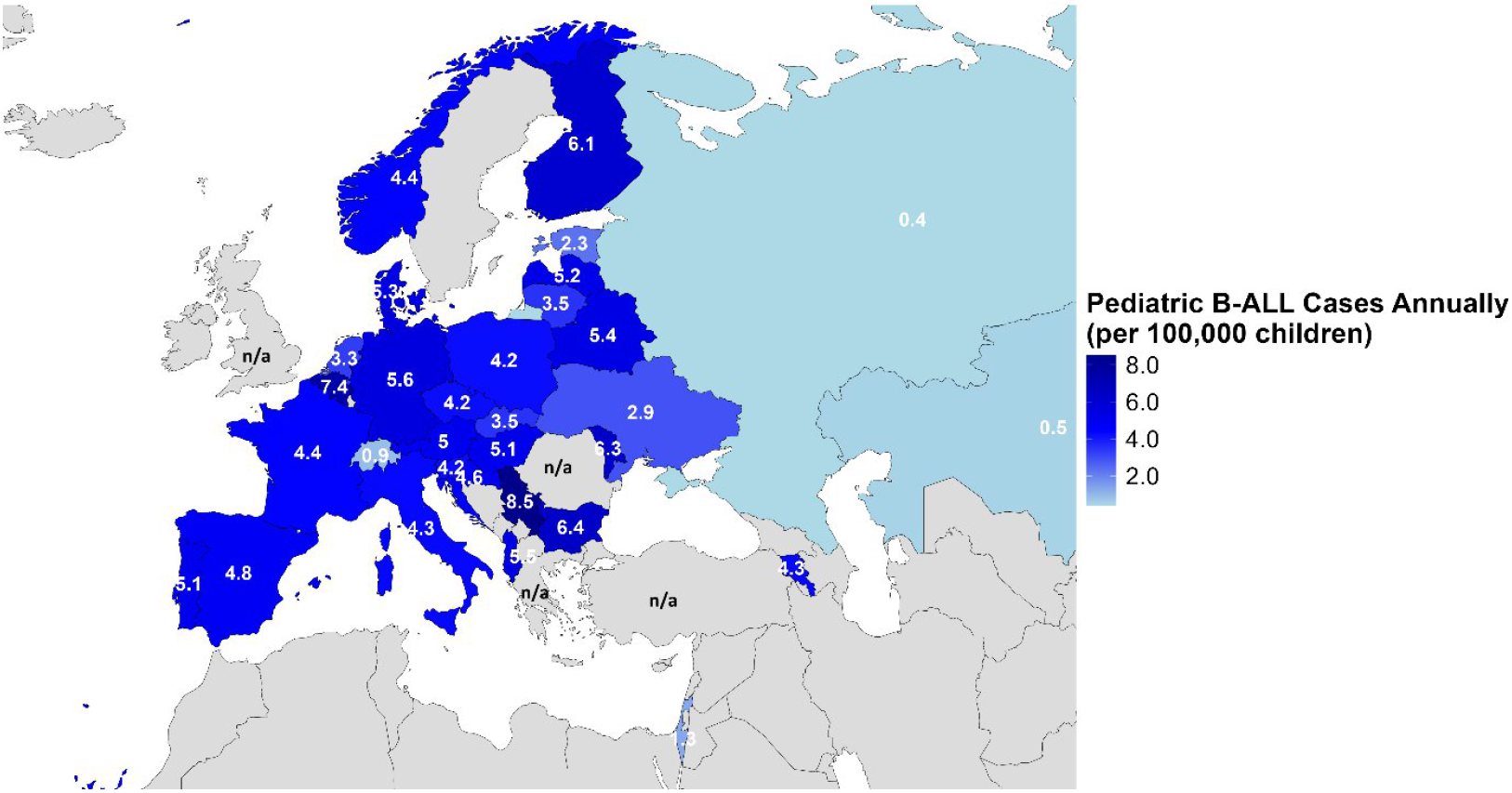
Availability of PHO centers and incidence of pediatric B-ALL across countries (A) Distribution of PHO centers per 1 million inhabitants. (B) Mean annual incidence of B-ALL per 100,000 children.

**Table 1.**
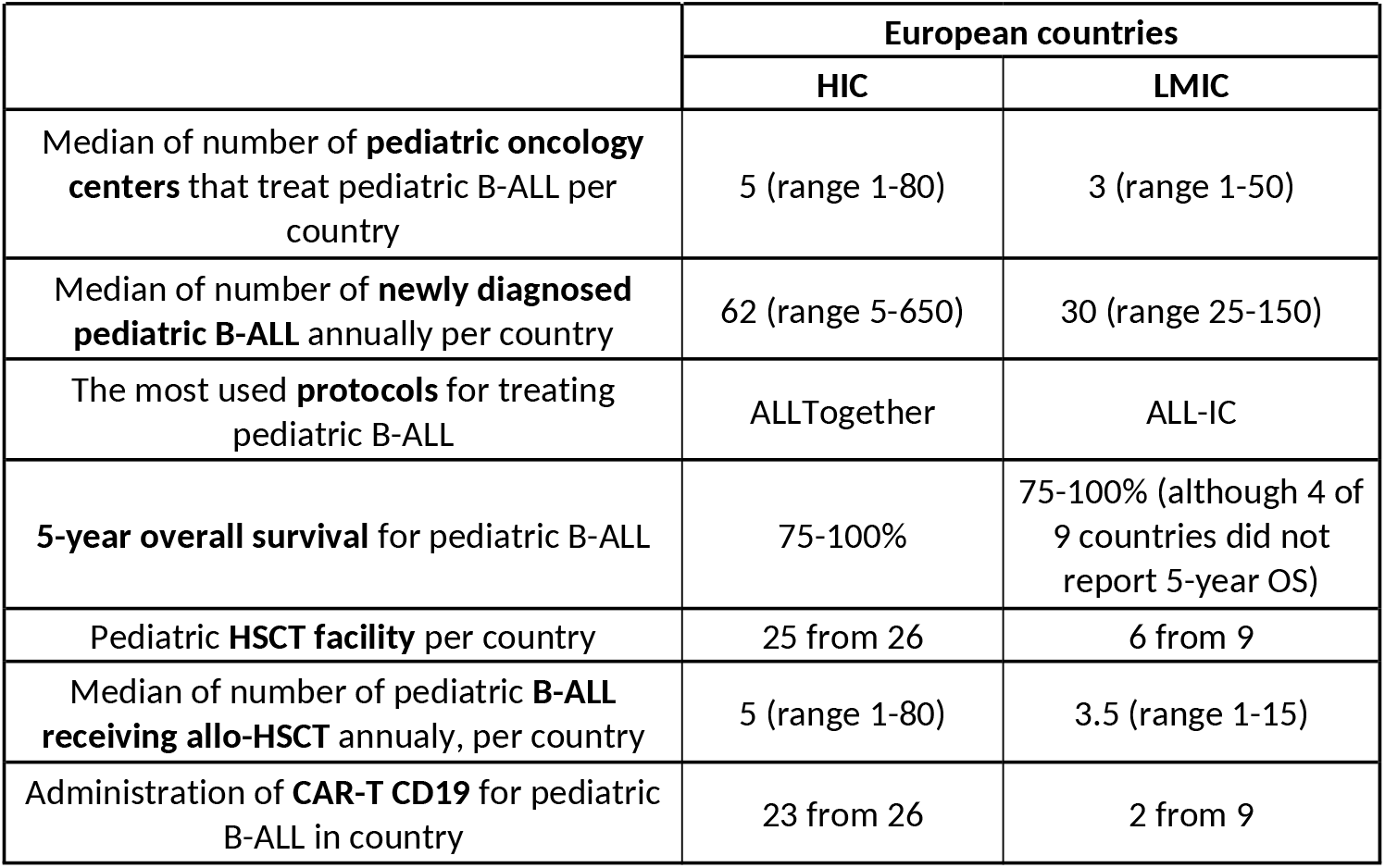
Comparison of resources and outcomes for pediatric B-ALL between HIC and LMIC European countries.

### Access to B-ALL therapy

The most commonly used treatment protocols were ALLTogether (12 countries), AIEOP-BFM ALL 2017 (9 countries), and ALL-IC (8 countries). In LMICs, ALL-IC was most frequently applied, although many respondents reported that they were not currently active participants in this research protocol. MRD assessment was limited in Moldova, Kazakhstan, and Albania, where only 0–25% of centers reported routine MRD testing, while Turkey reported 50–75% coverage. In HICs, MRD testing was consistently government-funded, whereas in LMICs, it was never government-funded in Albania, Kazakhstan, or Moldova.

Access to blinatumomab also varied in LMICs, it was unavailable in Moldova and Albania, limited (0–25% of eligible children) in Kazakhstan and Ukraine, and widely available in Armenia, Romania, Serbia, Turkey, and Belarus. In HICs, asparaginase was consistently government-funded, however, blinatumomab funding was variable. Spain, France, Italy, and the Netherlands reported only occasional funding for blinatumomab, and Switzerland reported it was rarely funded, depending on indication, with limited coverage for frontline disease. In LMICs, asparaginase was generally government-funded, whereas blinatumomab was never government-funded in Albania, Kazakhstan, Moldova or Ukraine.

### B-ALL survival

The reported 5-year overall survival (OS) for pediatric B-ALL exceeded 75% in most LMIC and all HICs, reaching above 90% in several HICs. OS data were unknown in Albania, Kazakhstan, Ukraine, and Moldova. Relapse rates were <25% in all HICs. In LMICs, 6 (66%) reported relapse rate <25%, with 2 countries (Armenia and Ukraine) reporting 25-50%, and one (Albania) relapse rate was unknown.Data sources for OS and relapse rates included nine national registries, 14 institutional records, and national B-ALL coordinator estimates from other countries.

### Hematopoietic stem cell transplantation (HSCT)

HSCT facilities were available in 89% of countries (n=31). Kazakhstan, Moldova, Albania, and Latvia reported no pediatric HSCT facilities. Among countries with HSCT access, a median of 5 children with B-ALL underwent transplantation per year (range: 1–80; Figure 3A). When stratified by income level, HICs reported a median of 5 transplants annually per country, with highest reported numbers in Italy (80 procedures per year). LMICs reported a median of 3.5 transplants annually per country.

**Figure 3.**
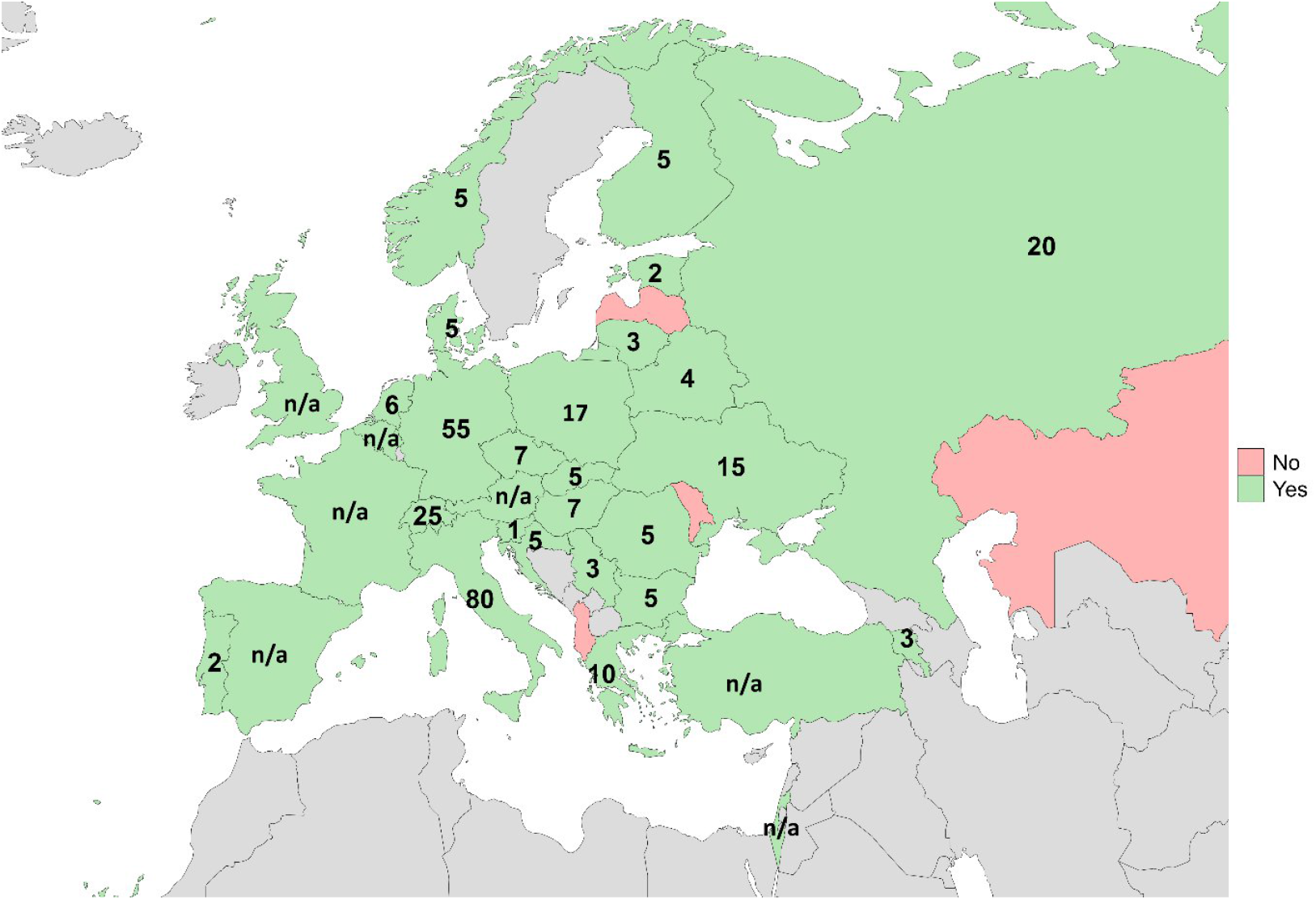
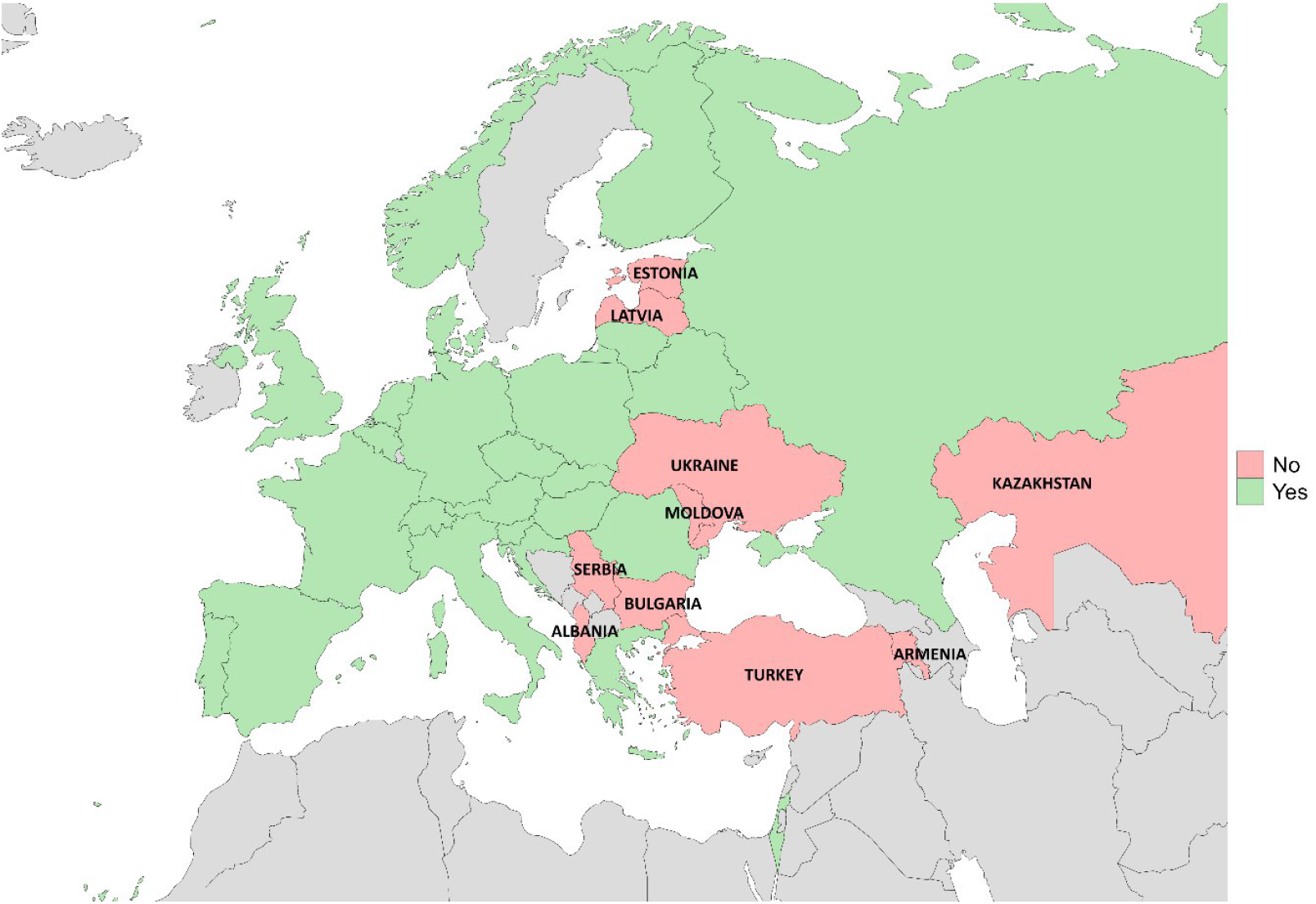
Availability of advanced therapies for pediatric B-ALL in Europe (A) Access to pediatric HSCT and average of children with B-ALL undergoing transplantation annually. (B) Access to CAR-T CD19 therapy.

In HICs, HSCT was accessible to nearly all eligible children, with Portugal being the only exception, where availability was limited to 50–75% of eligible cases. Seven countries (Estonia, Croatia, Israel, Latvia, Portugal, Bulgaria, and Russia) reported a perceived need to further expand HSCT access.

Four LMICs lacked transplant facilities entirely, while in the remaining five countries, HSCT was almost always available. However, Armenia reported that only 0–25% of eligible children underwent transplantation. Six of nine LMICs strongly agreed that HSCT access needed to be expanded, whereas the other three strongly disagreed.

In HICs, HSCT was consistently government-funded, whereas in LMICs, it was not funded in Albania, Kazakhstan and Moldova.

### CAR-T CD19 therapy

Across the region, CAR-T CD19 therapy for pediatric B-ALL was not administered in 10 countries: Armenia, Estonia, Kazakhstan, Latvia, Moldova, Ukraine, Albania, Serbia, Turkey, and Bulgaria.Overall, CAR-T CD19 therapy was available in 71% of assessed European countries that combined account for 80% of the European population (Figure 3B).

Among the 25 countries where CAR-T CD19 is available, the most common source was commercially purchased Novartis product. However, in Lithuania and Belarus, Tisagenlecleucel (Kymriah, Novartis) was not registered for pediatric use, and no availability was anticipated in the upcoming year. Nine countries reported having Point of Care CAR T-cell manufacturing facilities: Germany, Denmark, Spain, Finland, Greece, Italy, Lithuania, Belarus, and Russia. Across these 25 countries, 22 reported that 75–100% of eligible pediatric B-ALL patients had access to and received CAR-T CD19 therapy.Lower coverage (<75%) was reported in Italy, Belgium, and Switzerland.

More than half of surveyed countries did not have open clinical trials for CAR-T CD19. One country had trials exclusively for lymphoma, while 10 countries conducted trials for B-ALL or both B-ALL and lymphoma. Despite these limitations, CAR-T CD19 therapy was almost always or always funded by government sources. The most common eligibility criteria included refractory disease, second or subsequent relapse, or relapse after HSCT (Supplementary Table 1). The most frequently reported contraindications to CAR-T therapy were pregnancy, active HIV, HBV, or HCV infection, and chronic GVHD stage >II (Supplementary Table 2).

Exploring the availability of CAR-T through referral networks, 10 countries reported almost always or always accepting such patients, 10 accepted patients sometimes, three rarely, and Russia did not accept any. Inadequate funding was cited by 14 countries as the primary reason for declining international patients. On average, only 1–2 international patients were treated annually per country, while Spain and Belarus reported treating 20, and 10 international patients per year. While 17 countries expressed interest in participating in a referral network for patients from centers without CAR-T CD19 access, only six have established mechanisms for domestic or international referrals. Six countries were exploring international referral options, and eight were not interested in developing referral options. Funding for international treatment was most commonly provided by foundations, patients/families, or the referring country.

Among the 10 countries that did not administer CAR-T CD19 therapy domestically, six were able to refer patients abroad using existing networks. Mechanisms for international referral included [Supporting Action for Emergency Response](https://global.stjude.org/) (SAFER) Ukraine and the Princess Máxima Center network; Serbia referred patients to Italy and Austria. On average, Armenia, Albania, Turkey and Bulgaria reported referring 2–3 patients per year, with Ukraine reporting six patients annually through the SAFER Ukraine initiative. Funding was typically covered by patients or foundations.

For non-European Medicines Agency (non-EMA) approved therapies, 14 countries could send patients abroad. Specifically, four countries without domestic CAR-T access, Armenia, Estonia, Kazakhstan, and Ukraine could refer patients for non-EMA therapies. Serbia, Moldova, Turkey, and Latvia could only refer for EMA-approved therapies, while the remaining countries without CAR-T access were uncertain regarding non-EMA referrals (Supplementary Table 3).

### Barriers to referral network participation

Countries reported a range of factors limiting participation in CAR-T referral networks (Table 2). Financial barriers, including high therapy costs and limited funding or insurance coverage, were the most frequently cited, reported by 16 countries (46%). Regulatory challenges, such as national approvals, certification requirements, and ethical or legal considerations were identified by 6 countries (17%). Geographic limitations, including limited local CAR-T centers and logistical issues in transporting patient cells, were reported by 2 countries (6%). Small patient populations or limited experience were highlighted by 2 countries (6%). In contrast, 2 countries (6%), Spain and the United Kingdom, reported minimal or no barriers due to established CAR-T capacity and financing.Collaborative frameworks and cross-border arrangements were noted to facilitate access in Lithuania and Slovenia (2 countries, 6%).

**Table 2.** Barriers to CAR-T referral network participation by country.

| Barrier Type | Number of Countries | % of Reporting Countries | Countries Reporting |
| --- | --- | --- | --- |
| Financial barriers (high costs, limited funding/insurance) | 16 | 46% | Armenia, Albania, Austria, Belgium, Estonia, Hungary, Israel, Italy, Latvia, Moldova, Netherlands, Norway, Slovakia, Portugal, Turkey, Bulgaria |
| Regulatory challenges (approvals, certification, legal issues) | 6 | 17% | Albania, Switzerland, Finland, Turkey, Belarus, Kazakhstan |
| Geographic limitations (few CAR-T centers, logistics) | 2 | 6% | Kazakhstan, Lithuania |
| Small patient populations or limited experience | 2 | 6% | Czech Republic, Lithuania |
| No or minimal barriers | 2 | 6% | Spain, United Kingdom |
| Facilitated access through collaboration/cross-border networks | 2 | 6% | Lithuania, Slovenia |

## Discussion

This project aimed to assess access to CAR-T CD19 therapy and identify common barriers impacting its implementation for pediatric B-ALL across Europe. Our findings indicate that while most HICs provide domestic access to CAR-T therapy funded by the respective governments, many LMICs lack domestic CAR-T programs, rely on international referrals, and face financial or regulatory barriers. To fully understand the CAR-T landscape, we report both access to this therapy and information on the landscape of treatment of pediatric B-ALL. Our findings suggest that access to and referral pathways for CAR-T therapies remain variable throughout the region, with some countries facilitating access abroad and others facing restrictions or uncertainty. This highlights a unique opportunity to expand CAR-T CD19 access to benefit both high-income and low- and middle-income countries.

High-income countries reported a median of 62 new pediatric B-ALL cases annually, compared with 30 cases in LMICs. The lower numbers in LMICs likely reflect underdiagnosis rather than a true difference in incidence.^10^ The reported median of 5 pediatric hematology-oncology centers per country seems adequate to address the need for B-ALL care, as access to childhood cancer services should be within approximately 1–2 hours’ drive from a patient’s home.^11^ Proximity to care is crucial to address treatment-related side effects such as neutropenic fever, which requires antibiotic administration within the “golden hour” from the start of fever.^12^ In LMICs, where childhood cancer resources are more scarce, establishing dedicated and collaborative pediatric hematology-oncology centers, including those able to provide advanced therapy, may be the most practical and feasible way to ensure access to high-quality care for children with cancer.^13-15^

Many respondents from LMICs indicated that they were not active participants in international pediatric B-ALL protocols, and in some cases were using adapted protocols.^16^ Such adaptations are often necessary when access to essential diagnostic tools, such as minimal residual disease (MRD) monitoring^17^, is lacking, as reported by Moldova, Kazakhstan, and Albania. Moreover, limited supportive care resources, such as capacity for infection management, transfusion support, and intensive care, also necessitate local protocol modifications, as not all centers are equipped to manage treatment-related toxicities.^18^ Interestingly, although MRD testing was not consistently funded by governments in LMICs, asparaginase was almost always available, which is a positive finding given its crucial role in B-ALL regimens.^19^

In addition to access to CAR-T CD19, we collected information on accessibility of other advanced therapies, including blinatumomab and HSCT. Half of LMICs reported limited or no access to blinatumomab, with no funding available, whereas some of these countries reported access to HSCT. This discrepancy is concerning, as blinatumomab significantly enhances event-free-survival and reduces toxicity in relapsed patients.^20^ Blinatumomab is approved by the EMA for relapsed or refractory disease, but not for first-line use in pediatric patients. Even in high-income countries, access to blinatumomab is often constrained by funding policies, with use outside the approved indication typically requiring case-by-case health insurance authorization. Extended pediatric approval is needed to facilitate use in upfront therapy, where it has been shown to improve disease-free survival.^21^

For patients who relapse after HSCT, have second or further relapse, or leukemia refractory to treatment, CAR-T CD19 remains the last therapeutic option.^22^ Yet CAR-T CD19 therapy for pediatric B-ALL is not available in one third (32%) of European countries. Where access does exist, tisagenlecleucel^6^ was most commonly used. Only nine of 25 countries reported having a local CAR-T manufacturing facility. Point-of-care manufacturing represents a promising strategy to universalize CAR-T cell therapy by bringing the production process closer to the patient, thereby reducing costs, shortening production times, and improving accessibility, particularly in underserved regions.^23-25^ Development of additional CAR-T production pathways remains an essential gap to reduce the cost of access to CAR-T in the region.^7,26^ Additionally, more than half of countries reported no open CAR-T clinical trials, presenting another opportunity to reduce costs and strengthen scientific collaboration in this field.^27^

Importantly, two-thirds of countries expressed willingness to participate in a referral network to admit children for CAR-T from abroad. At present, only two patients per country per year receive CAR-T therapy internationally and only six countries have established mechanisms to send patients abroad, often relying on existing programs such as SAFER Ukraine^28^ or the Princess Máxima Center network. These mechanisms are frequently supported by foundations covering treatment costs, thus making access often contingent on variable funding availability.^29^ Supportive policies and cross-border agreements are essential to ensure equitable access beyond EMA-approved options.^30^ Notably, half of the countries without CAR-T CD19 access are able to send patients abroad for non-EMA-approved therapy, underlining the importance of such mechanisms in creating opportunities for referral pathways.

Barriers to implementing CAR-T reflect a mix of financial, regulatory, and geographic challenges, with considerable country-specific variation tied to differences in healthcare infrastructure and policy.

While established referral pathways in some countries minimize obstacles, others must rely on collaborative or cross-border networks to address gaps in capacity and expertise. The result is a heterogeneous landscape across Europe, in which financial, regulatory, and logistical considerations variably shape CAR-T access for patients.

This study has several limitations. Very small countries (e.g., Andorra, Iceland, Luxembourg, Malta, Monaco, and San Marino) were not included in the study due to the absence of established contacts. Among the remaining 43 countries, 7 did not respond (Azerbaijan, Ireland, Kyrgyzstan, Sweden, Tajikistan, Turkmenistan, and Uzbekistan), resulting in a coverage of 36 out of 43 countries (84%).

Wherever possible, we sought to obtain responses from national coordinators to reflect a country-level perspective. A national coordinator refers to the physician responsible for leading and coordinating the treatment of pediatric B-ALL within their country, including the oversight of national protocols and treatment standards. However, in countries without a designated national coordinator for pediatric B-ALL, we relied on institutional responses. In some cases, data from a few institutions were used to approximate a national perspective. This approach introduces potential bias, as institutional practices may not fully represent country-wide reality. Additionally, in certain instances, the responses were based primarily on the respondents’ expert opinions rather than systematically collected or documented national statistics. This further underscores the need for comprehensive registries and standardized documentation systems to ensure more accurate and representative data in future assessments.

To our knowledge, this is the first multi-country evaluation of CAR-T CD19 access mapping in pediatric B-ALL in Europe. Building on this foundation, micro-costing and cost-effectiveness analyses will be conducted to generate real-world evidence on the costs associated with CAR-T and its alternative treatment options, thereby the development of additional strategies for equitable CAR-T implementation in low- and middle-income countries. In subsequent phases, the project will expand beyond Europe to other underserved regions, including Latin America and the Mediterranean region.

This initiative enhances understanding of demand for CAR-T CD19 and identifies barriers to access in Europe. Its broader goal is to reduce global disparities in pediatric leukemia survival by developing scalable, evidence-based models for equitable access to advanced therapies such as CAR-T. Despite the European Union’s strong focus on rare diseases, no standardized pathway for CAR-T referral currently exists and access remains variable across the region. A coordinated regional framework is urgently needed to promote equitable access, sustainable funding, and patient referral coordination. The findings of this project may inform the development of such a framework and contribute to the long-term goal of ensuring that every child, regardless of geography, can access potentially curative treatment.

## Declarations

### Ethics approval and consent to participate

The study was conducted in accordance with ethical principles for observational research. Approval was obtained from the Bioethics Committee at the Medical University of Lodz, Poland (RNN/56/24/KE) and from the St Jude Children’s Research Hospital, USA (IRB number: 24-1650).Participation was voluntary, no individual patient identifiers were collected, and data were analyzed in aggregate to maintain confidentiality.

### Data-sharing statement

Data are available upon request to the corresponding author.

### Authors’ contributions

Conceptualization – AO, APM, NB, JS, CRG, WM, AA, CD, KK, KS

Data curation – AO, JEG, AD, JW, MD, SJ, CD, KK, KS

Formal analysis - AO, JW, MD

Funding acquisition - AO

Investigation – AO, JEG, AD, JW, MD, APM, SJ, NB, JS, TY, CRG, WM, AA, CD, KK, KS

Methodology – AO, JEG, AD, JW, MD, AA, CD, KK, KS

Project administration – AO, JEG, AD

Resources – AO, JEG, AD, AA, KK, KS

Software – AO, JW, MD, AA

Supervision –JS, CRC, WM, AA, CD, KK, KS

Validation - AO, JEG, AD, JW, MD, APM, NB, SJ, JS, TY, CRC, WM, AA, CD, KK, KS

Visualization - AO

Writing – original draft - AO, KS

Writing – review & editing - AO, JEG, AD, JW, MD, APM, NB, SJ, JS, TY, CRG, WM, AA, CD, KK, KS

## Data Availability

Data are available upon request to the corresponding author.

## Acknowledgement

We gratefully acknowledge the collaborative contributions of St. Jude Global, the European Group for Blood and Marrow Transplantation (EBMT) Pediatric Disease Working Party (PDWP), and the International BFM Study Group (I-BFM), as well as the pediatric oncology network. We also extend our sincere thanks to colleagues at participating institutions for their valuable input and support.

