## Supplements for "Landscape assessment to characterize baseline access and multilevel barriers to IMProve Access to CAR-T CD19 therapy (IMPACT study) across Europe"

**Supplement 1**

### IMPACT- CAR-T Cell Institutional Assessment Questionnaire

#### GENERAL INFORMATION

1. Full Name ______________________________________________
2. Email Address ______________________________________________
3. Current Place of Work ______________________________________________

4. In which country do you currently reside? ________________________

5. Do you currently serve as a National Coordinator for pediatric B-cell precursor acute lymphoblastic leukemia (BCP-ALL) treatment?

- Yes
- No

If not please list the name of the national coordinator or enter "unknown":________________

6. Please select your current position(s) in the pediatric oncology field in your country (select all that apply):

- Head of Department of pediatric oncology and hematology
- Member of European Society for Blood and Marrow Transplantation (EBMT)
- Specialist in pediatric oncology and hematology
- Transplantologist
- Medical doctor
- Other

If other, please specify __________________________________________________

#### COUNTRY LEVEL CAPACITY

7. How many Pediatric Onco-Hematology Departments or treatment facilities are in your **country**?___________________________________________

8. How many departments or treatment facilities in your **country** treat children with B-cell precursor acute lymphoblastic leukemia (BCP-ALL)?________________________________

9. Does your **country** have departments or treatment facilities that perform hematopoietic stem cell transplantation (HSCT) for **children** with BCP-ALL?

- Yes
- No

10. Pease provide how many **adult** centers are certified for HSCT in your **country:**__________

11. How many departments or treatment facilities are certified for hematopoietic stem cell transplantation (HSCT) for children with BCP-ALL in your **country**?_____________________

12. Based on your experience, indicate the degree to which you agree with the following statement. There is a need to increase access to hematopoietic stem cell transplantation (HSCT) procedures for children with BCP-ALL in my **country**.
 * Access = affordable and was administered at the time it was needed

- Strongly disagree
- Disagree
- Neither agree nor disagree
- Agree
- Strongly agree

13. In your **country**, what proportion of centers measure minimal residual disease (MRD) and use it to guide therapy for children with BCP-ALL?

- 0 - 25%
- 25 - 50%
- 50 - 75%
- 75 - 100%

14. **How often are the following diagnostic modalities or treatments funded by the government (available and provided at no cost to the patients)?**

| | Always | Almost always | Sometimes | Rarely | Never | Unknown |
|---|---|---|---|---|---|---|
| Minimal residual disease (MRD) for children with BCP- ALL | | | | | | |
| Asparaginase for children with BCP- ALL | | | | | | |
| Blinatumomab for children with BCP- ALL | | | | | | |
| Allogeneic hematopoietic stem cell transplantation (HSCT) for children with BCP- ALL | | | | | | |

If not always available, please provide additional comments:___________________________

**ACUTE LYMPHOBLASTIC LEUKEMIA**

15. Approximately how many children (age 0-18) on average are diagnosed with BCP-ALL in your **institution** per year (n per year)?
*(Enter 0 if none, enter 9999 if you don't have access to this data or you don't have access to this information)*_________________________________________________

16. What is the overall survival rate (OS) of childhood BCP-ALL in your **institution** (% of treated children – age 0-18)?

- 0 - 25%
- 25 - 50%
- 50 -75%
- 75 - 100%
- Unknown

17. Timepoint of overall survival estimate:

- 1 year
- 2 years
- 3 years
- 4 years
- 5 years
- 6 years
- Other

If other, please specify:____________________________________________________

18. What is the relapse rate (number of patients that relapsed out of 100 patients) of childhood (age 0-18) BCP-ALL in your institution?

- 0 - 25%
- 25 - 50%
- 50 - 75%
- 75 - 100%
- Unknown

19. What was the source for the data provided?

- National registry
- Last ALL study conducted at the institution
- Institutional Records
- Estimate
- Other

If other, please specify the source for the data provided:________________________________

20. Is your **institution** participating in any of the following active ALL collaborative group protocols?

- AIEOP-BFM ALL 2017
- ALL-IC BFM
- ALLTogether
- No, we do not participate in any of these current ALL protocols

21. Is minimal residual disease (MRD) testing a part of the standard of care for BCP-ALL at your **institution**?

- Yes
- No

22. What proportion of children diagnosed with BCP-ALL have MRD testing as part of their treatment regimen?

- 0 - 25%
- 25 - 50%
- 50 - 75%
- 75 - 100%
- Unknown

23. Which MRD technique(s) are used in your **institution**? (select all that apply)

- Flow-MRD
- NGS-MRD
- PCR-MRD
- None
- Other

If other, please specify:_____________

24. Please indicate the availability* of Asparaginase children with BCP-ALL at your **institution:**
** Available = commercially available and either in hospital or can be ordered and ready for administration in a timely manner.*

- Always
- Almost always
- Sometimes
- Rarely
- Never

25. Of the children with BCP-ALL in your **institution**, what proportion of patients miss a dose of Asparaginase?

- 0 - 25%

- 25 - 50%
- 50 - 75%
- 75 - 100%
- Unknown

26. Which preparation(s) of Asparaginase are used in your **institution?** (select all that apply)

- L-Asparaginase
- PEG-asparaginase
- Erwinia
- Other

If other, please specify:____________________________________

**TREATMENT AVAILABILITY**

27. Please indicate the availability * of blinatumomab for children with BCP-ALL at your **institution:**

**Available = commercially available and either in a hospital or can be ordered and ready for administration in a timely manner*

- Always
- Almost always
- Sometimes
- Rarely
- Never

**28.** What proportion of eligible children have access* to blinatumomab as part of their BCP-ALL treatment?

** Access = affordable and was administered at time it was needed*

- 0 - 25%
- 25 - 50%
- 50 -75%
- 75 - 100%
- Unknown

29. How many children with BCP-ALL receive blinatumomab in your **institution** annually (n per year)?

*(Enter 0 if none, enter 9999 if you don't have access to this data or you don't have access to this information)*_________________________________________

30. Please indicate the availability* of allogeneic hematopoietic stem cell transplantation (HSCT) for children with BCP-ALL at your institution:

**Available = commercially available and either in hospital or can be ordered and ready for administration in a timely manner.*

- Always
- Almost always
- Sometimes
- Rarely
- Never

31. What proportion of eligible (determined based on institution criteria) children have access to HSCT as part of their BCP-ALL treatment?
** Access = able to afford and was administered at time it was needed*
- 0 - 25%
- 25 - 50%
- 50 -75%
- 75 - 100%
- Unknown

32. How many children with BCP-ALL receive HSCT in your **institution** annually (n per year)? *(Enter 0 if none, enter 9999 if you don't have access to this data or you don't have access to this information)*_______________________________________________________

**CAR-T CD19 CELLS THERAPY**

33. CAR-T cells administered at your institution?
- Yes
- No

34. What is the source of CAR-T CD19 cells products at your **institution**? (select all that apply)
- CAR-T cells manufacturing facility in my institution (clinical academic production facility)
- CAR-T cells manufacturing facility at a neighboring institution in my country
- CAR-T cells accessible through international collaborative research protocols
- Purchase pre-made CAR-T cells from a manufacturing facility in a neighboring country (not an academic production center)
- Buying commercially available pre-made CAR-T cells from Novartis
- Other

If other, please specify:____________________________________________________________

35. What proportion of patients with BCP-ALL eligible for CAR-T cells have access to and receive them?
- 0 - 25%
- 25 - 50%
- 50 - 75%
- 75 - 100%
- Unknown

36. Are there open clinical trials or international collaboration for children to receive CAR-T cells therapy (for BCP-ALL, or lymphoma) in your **institution**?

- Yes for both BCP-ALL and lymphoma
- Yes, but only for BCP-ALL
- Yes, but only for lymphoma
- No
- Unknown

37. Please write names or provide the website with information about active or future clinical trials or international collaborations for CAR-T CD19 cell therapy for children in your **institution**:_______________________________________________________________

**38.** How often is CAR-T CD19 cell therapy fully covered reimbursed by the government **(in your country)**?

- Always
- Almost always
- Sometimes
- Rarely
- Never

**39.** Please choose what kind of sources of funding CAR-T CD19 cells therapy for children with BCP-ALL are available in your **country** (select all that apply).

- Patient/family
- Crowdfunding/crowdsourcing
- Local foundation/NGO
- International NGO
- Other

If other, please specify:_______________________________________________________

40. Is **Tisagenlecleucel** (Kymriah, Novartis) registered for children in your country?

- Yes
- No, do not anticipate it will be available by next year
- No, but in process (expect it to be available within 1 year)
- Unknown

41. What are the eligibility criteria for CAR-T CD19 therapy for children with BCP-ALL in your **institution**? (select all that apply)

- Failure to achieve complete remission (CR) after 2 cycles of standard chemotherapy (primary resistance for treatment)
- Failure to achieve CR after one cycle of reinduction therapy for relapsed ALL (secondary resistance to treatment)

- Second or subsequent recurrence in the bone marrow (disease recurrence)
- Relapse (in bone marrow) after allogeneic hematopoietic stem cell transplantation (allo-HSCT) and after an interval of at least 4 months between allo-HSCT and CAR-T administration
- Failure of at least two lines of tyrosine kinase inhibitor (TKI) therapy or, in the case of the EsPhALL2017 protocol, failure to achieve complete remission (CR) after consolidation. Indications also include intolerance or contraindications to TKI therapy
- Allo-HSCT cannot be performed (Possible causes: comorbidities, contraindications to conditioning treatment before allo-HSCT, lack of a suitable donor (e.g. only available haploidentical donor with indications only for a compatible donor according to the protocol) or previous allo-HSCT)
- Children above 6 months old
- Children less than 18 years old
- Other

If other, please specify other criteria:_______________________________________________

42. What are the contraindications for CAR-T CD19 therapy for children with BCP-ALL in your **institution**? Please select all that apply.

- Pregnancy or breastfeeding
- Isolated extramedullary ALL recurrence
- Central nervous system involvement by ALL (at the time of administration of CAR-T CD19)
- Actively uncontrolled systemic infection
- HIV infection
- Active hepatitis B or C
- Coexistence of congenital genetic diseases with impaired bone marrow function (e.g. Fanconi anemia, Kostmann syndrome, Shwachman-Diamond syndrome)
- Chronic graft-versus-host disease (cGvHD) stage II-IV
- Presence of contraindications to the use of lymphodepleting chemotherapy (i.e. fludarabine and cyclophosphamide or cytarabine and etoposide)
- Age over 18 years old
- Other

If other, please specify other contraindications:________________________________________

43. Does your **institution** accept patients from external centers for CAR-T cell therapy?

- Always
- Almost always
- Sometimes
- Rarely
- Never

44. How often is inadequate funding the primary reason why international patients are not accepted?

- Always
- Almost always
- Sometimes
- Rarely
- Never

45. How many children with BCP-ALL **from another country** receive CAR-T CD19 therapy in your **institution** annually?
*(Enter 0 if none, enter 9999 if you don't have access to this data or you don't have access to this information)*______________________________________________________

46. What is the typical source of funding for CAR-T CD19 therapy for **international** patients in your **institution**? (select all that apply)

- Funding is covered by primary patient Institution
- Funding is covered by hosting/accepting Institution
- Funding is covered by a foundation based in the referring country
- Funding is covered by foundation in the accepting country
- Patients paid privately (complete funded by patient family)
- It is covered by government in hosting/accepting Institution
- Other
- Not Applicable

If other, please specify:_____________________________________________

47. Are you able to refer children for CAR-T cells at a different institution?

- Yes, refer to another center in my country
- Yes, refer to another center in a different country
- No, there is no current mechanism to refer patients for CAR-T cells
- Other

If other, please specify:____________________________________________

48. Would your **institution** be interested in participating in a referral network for patients from **institutions** without access to CAR-T CD19 cells therapy?

- Yes
- Maybe
- No
- Unknown

49. Does your institution have an established mechanism or partnership to send patients with BCP-ALL to another institution in your country or abroad for CAR-T CD19 cells therapy?

- Yes
- No

If yes, select all that apply
- Yes, in my country
- Yes, internationally

Please write to which institutions/countries:___________________________________

If no, select all that apply
- No, but we are looking for in country options
- No, but we are looking for international options
- No, we are not interested

50. How many children from your **institution** with BCP-ALL travel to another country for CAR-T CD19 therapy annually?
*(Enter 0 if none, enter 9999 if you don't have access to this data or you don't have access to this information)*
Independently/individual_______________________________________________________
Using established partnerships _________________________________________

**51.** Please select all funding sources available through your **institution** to support the treatment of patients with CAR-T cells therapy abroad. (select all that apply)
- Government funded
- Hospital funded
- Patient/family
- Crowd sourcing/funding
- Local Foundation/NGO
- International NGO
- Private insurance
- Other
- None

If other, please specify:__________________________________________________________

52. Is your **institution** legally allowed to send patients abroad from nonEMA (European Medicines Agency) approved therapy (such as an academic-based CAR-T product)?
- Yes
- No, only for EMA approved
- No
- Unknown
- Other

If other, please specify:_____________________________________________

53. Please provide details about which factors (financial, regulatory, geographic, …) impact your institutions ability to participate in a referral network for CAR-T CD19?___________________

If you selected 9999 because you do not have access to data or information to answer a particular question, please provide contact information (email address) for a person with access to this data or type "Unknown" if there is no one with access to this information:____________

Please list the names of the people who contributed or collected to the data for this survey:____

Please add any additional comments related to  CAR-T CD19 cell therapy or the questionnaire below.______________________________________________________________

Thank you so much for filling this questionnaire and participating in our study!

**Supplement 2**

#### IMPACT- CAR-T Cell Country Assessment Questionnaire

Dear all,

You have been recognized as a pediatric oncology and hematology collaborator and leader in your country. Together with The European Group for Blood and Marrow Transplantation (EBMT) – Pediatric Disease Working party, International BFM Study Group, Committee ALL and St. Jude Children's Research Hospital we would like to invite you to participate in IMPACT project (Improve Access to CAR-T CD19 therapy in European region).

We kindly ask you to spend a few minutes on this survey, which will allow us to better understand access to advanced therapy (CAR-T CD19 cells) in European countries. Feel free to share it with your colleagues, if you think they will contribute to this study.

Thank you!

##### GENERAL INFORMATION

1. Full Name ____________________________________________________
2. Email Address ____________________________________________________
3. Current Place of Work ____________________________________________________

4. In which country do you currently reside? __________________________

5. Do you currently serve as a **National Coordinator** for pediatric B-cell precursor acute lymphoblastic leukemia (BCP-ALL) treatment?

- Yes
- No

If not please list the name of the national coordinator or enter "unknown":_________________

6. Please select your current position(s) in the pediatric oncology field in your country (select all that apply):

- Head of Department of pediatric oncology and hematology
- Member of European Society for Blood and Marrow Transplantation (EBMT)
- Specialist in pediatric oncology and hematology
- Transplantologist
- Medical doctor
- Other

If other, please specify:
________________________________________________________________

#### COUNTRY LEVEL CAPACITY

6. How many Pediatric Onco-Hematology Departments or treatment facilities are in your **country**?_______________________________________________

7. How many departments or treatment facilities in your **country** treat children with B-cell precursor acute lymphoblastic leukemia (BCP-ALL)?________________

8. Does your **country** have departments or treatment facilities that perform hematopoietic stem cell transplantation (HSCT) for **children** with BCP-ALL?

- Yes
- No

9. Pease provide how many **adult** centers are certified for HSCT in your **country:**__

10. How many departments or treatment facilities are certified for hematopoietic stem cell transplantation (HSCT) for children with BCP-ALL in your **country**?_________

11. Based on your experience, indicate the degree to which you agree with the following statement. There is a need to increase access to hematopoietic stem cell transplantation (HSCT) procedures for children with BCP-ALL in my **country**.
 * Access = affordable and was administered at the time it was needed

- Strongly disagree
- Disagree
- Neither agree nor disagree
- Agree
- Strongly agree

12. In your **country**, what proportion of centers measure minimal residual disease (MRD) and use it to guide therapy for children with BCP-ALL?

- 0 - 25%
- 25 - 50%
- 50 - 75%
- 75 - 100%

13. **How often are the following diagnostic modalities or treatments funded by the government (available and provided at no cost to the patients)?**

| | Always | Almost always | Sometimes | Rarely | Never | Unknown |
|---|---|---|---|---|---|---|
| Minimal residual | | | | | | |

| |
|---|
| disease (MRD) for children with BCP- ALL |
| Asparaginase for children with BCP- ALL |
| Blinatumomab for children with BCP- ALL |
| Allogeneic hematopoietic stem cell transplantation (HSCT) for children with BCP- ALL |

If not always available, please provide additional comments:___________________________

**ACUTE LYMPHOBLASTIC LEUKEMIA**

14. Approximately how many children (age 0-18) on average are diagnosed with BCP-ALL in your **country** per year (n per year)?
 *(Enter 0 if none, enter 9999 if you don't have access to this data or you don't have access to this information)*________________________________

15. What is the overall survival rate (OS) of childhood BCP-ALL in your **country** (% of treated children – age 0-18)?

- 0 - 25%
- 25 - 50%
- 50 -75%
- 75 - 100%
- Unknown

16. Timepoint of overall survival estimate:

- 1 year
- 2 years
- 3 years
- 4 years
- 5 years
- 6 years
- Other

17. What is the relapse rate (number of patients that relapsed out of 100 patients) of childhood (age 0-18) BCP-ALL in your **country**?

- 0 - 25%
- 25 - 50%
- 50 - 75%
- 75 - 100%
- Unknown

18. What was the source for the data provided?

- National registry
- Last ALL study conducted at the institution
- Institutional Records
- Estimate
- Other

If other, please specify the source for the data provided:______________________________

19. Is your **country** participating in any of the following active ALL collaborative group protocols?

- AIEOP-BFM ALL 2017
- ALL-IC BFM
- ALLTogether
- No, we do not participate in any of these current ALL protocols

20. Is minimal residual disease (MRD) testing a part of the standard of care for BCP-ALL in your **country**?

- Yes
- No

21. What proportion of children diagnosed with BCP-ALL have MRD testing as part of their treatment regimen?

- 0 - 25%
- 25 - 50%
- 50 - 75%
- 75 - 100%
- Unknown

22. Which MRD technique(s) are used in your **country**? (select all that apply)

- Flow-MRD
- NGS-MRD
- PCR-MRD
- None

- Other

If other, please specify:_______________________________________________

23. Please indicate the availability* of Asparaginase for children with BCP-ALL in your **country:**
** Available = commercially available and either in hospital or can be ordered and ready for administration in a timely manner.*

- Always
- Almost always
- Sometimes
- Rarely
- Never

24. Of the children with BCP-ALL in your **country**, what proportion of patients miss a dose of Asparaginase?

- 0 - 25%
- 25 - 50%
- 50 - 75%
- 75 - 100%
- Unknown

25. Which preparation(s) of Asparaginase are used in your **country?** (select all that apply)

- L-Asparaginase
- PEG-asparaginase
- Erwinia
- Other

If other, please specify:________________________________________

**TREATMENT AVAILABILITY**

26. Please indicate the availability * of blinatumomab for children with BCP-ALL in your **country:**
**Available = commercially available and either in a hospital or can be ordered and ready for administration in a timely manner*

- Always
- Almost always
- Sometimes
- Rarely
- Never

27. What proportion of eligible children have access* to blinatumomab as part of their BCP-ALL treatment?
** Access = affordable and was administered at time it was needed*

- 0 - 25%
- 25 - 50%
- 50 -75%
- 75 - 100%
- Unknown

28. How many children with BCP-ALL receive blinatumomab in your **country** annually (n per year)?
*(Enter 0 if none, enter 9999 if you don't have access to this data or you don't have access to this information)*____________________________________________

29. Please indicate the availability* of allogeneic hematopoietic stem cell transplantation (HSCT) for children with BCP-ALL in your **country**:
**Available = commercially available and either in hospital or can be ordered and ready for administration in a timely manner.*

- Always
- Almost always
- Sometimes
- Rarely
- Never

30. What proportion of eligible (determined based on country criteria) children have access to HSCT as part of their BCP-ALL treatment?
** Access = able to afford and was administered at time it was needed*

- 0 - 25%
- 25 - 50%
- 50 -75%
- 75 - 100%
- Unknown

31. How many children with BCP-ALL receive HSCT in your **country** annually (n per year)?
*(Enter 0 if none, enter 9999 if you don't have access to this data or you don't have access to this information)*______________________________________________

**CAR-T CD19 CELLS THERAPY**

32. Are CAR-T cells administered in your country?

- Yes
- No

33. What is the source of CAR-T CD19 cells products in your **country**? (select all that apply)

- CAR-T cells manufacturing facility in my country (at least one or more clinical academic production facility)
- CAR-T cells manufacturing facility at a neighboring institution in my country
- CAR-T cells accessible through international collaborative research protocols
- Purchase pre-made CAR-T cells from a manufacturing facility in a neighboring country (not an academic production center)
- Buying commercially available pre-made CAR-T cells from Novartis
- Other

If other, please specify:___________________________________________________

34. What proportion of patients with BCP-ALL eligible for CAR-T cells have access to and receive them?

- 0 - 25%
- 25 - 50%
- 50 - 75%
- 75 - 100%
- Unknown

35. Are there open clinical trials or international collaboration for children to receive CAR-T cells therapy (for BCP-ALL, or lymphoma) in your **country**?

- Yes for both BCP-ALL and lymphoma
- Yes, but only for BCP-ALL
- Yes, but only for lymphoma
- No
- Unknown

36. Please write names or provide the website with information about active or future clinical trials or international collaborations for CAR-T CD19 cell therapy for children in your **country**:__

37. How often is CAR-T CD19 cell therapy fully covered reimbursed by the government **(in your country)**?

- Always
- Almost always
- Sometimes
- Rarely
- Never

38. Please choose what kind of sources of funding CAR-T CD19 cells therapy for children with BCP-ALL are available in your **country** (select all that apply).

- Patient/family
- Crowdfunding/crowdsourcing

- Local foundation/NGO
- International NGO
- Other

If other, please specify:_______________________________________________

39. Is **Tisagenlecleucel** (Kymriah, Novartis) registered for children in your country?
    - Yes
    - No, do not anticipate it will be available by next year
    - No, but in process (expect it to be available within 1 year)
    - Unknown

40. What are the eligibility criteria for CAR-T CD19 therapy for children with BCP-ALL in your **country**? (select all that apply)
    - Failure to achieve complete remission (CR) after 2 cycles of standard chemotherapy (primary resistance for treatment)
    - Failure to achieve CR after one cycle of reinduction therapy for relapsed ALL (secondary resistance to treatment)
    - Second or subsequent recurrence in the bone marrow (disease recurrence)
    - Relapse (in bone marrow) after allogeneic hematopoietic stem cell transplantation (allo-HSCT) and after an interval of at least 4 months between allo-HSCT and CAR-T administration
    - Failure of at least two lines of tyrosine kinase inhibitor (TKI) therapy or, in the case of the EsPhALL2017 protocol, failure to achieve complete remission (CR) after consolidation. Indications also include intolerance or contraindications to TKI therapy
    - Allo-HSCT cannot be performed (Possible causes: comorbidities, contraindications to conditioning treatment before allo-HSCT, lack of a suitable donor (e.g. only available haploidentical donor with indications only for a compatible donor according to the protocol) or previous allo-HSCT)
    - Children above 6 months old
    - Children less than 18 years old
    - Other

If other, please specify other criteria:___________________________________

41. What are the contraindications for CAR-T CD19 therapy for children with BCP-ALL in your **country**? Please select all that apply.
    - Pregnancy or breastfeeding
    - Isolated extramedullary ALL recurrence
    - Central nervous system involvement by ALL (at the time of administration of CAR-T CD19)
    - Actively uncontrolled systemic infection
    - HIV infection

- Active hepatitis B or C
- Coexistence of congenital genetic diseases with impaired bone marrow function (e.g. Fanconi anemia, Kostmann syndrome, Shwachman-Diamond syndrome)
- Chronic graft-versus-host disease (cGvHD) stage II-IV
- Presence of contraindications to the use of lymphodepleting chemotherapy (i.e. fludarabine and cyclophosphamide or cytarabine and etoposide)
- Age over 18 years old
- Other

If other, please specify other contraindications:____________________________

42. Does your **country** accept patients from external centers for CAR-T cell therapy?
- Always
- Almost always
- Sometimes
- Rarely
- Never

43. How often is inadequate funding the primary reason why international patients are not accepted?
- Always
- Almost always
- Sometimes
- Rarely
- Never

44. How many children with BCP-ALL **from another country** receive CAR-T CD19 therapy in your **country** annually?
*(Enter 0 if none, enter 9999 if you don't have access to this data or you don't have access to this information)*____________________________________________

45. What is the typical source of funding for CAR-T CD19 therapy for **international** patients in your **country**? (select all that apply)
- Funding is covered by primary patient Institution
- Funding is covered by hosting/accepting Institution
- Funding is covered by a foundation based in the referring country
- Funding is covered by foundation in the accepting country
- Patients paid privately (complete funded by patient family)
- It is covered by government in hosting/accepting Institution
- Other
- Not Applicable

If other, please specify:_____________________________________________

46. Are you able to refer children for CAR-T cells at a different **country**?

- Yes, refer to another center in my country
- Yes, refer to another center in a different country
- No, there is no current mechanism to refer patients for CAR-T cells
- Other

If other, please specify:_____________________________________________________

47. Would your **country** be interested in participating in a referral network for patients from **institutions** without access to CAR-T CD19 cells therapy?

- Yes
- Maybe
- No
- Unknown

Please explain:_________________________________________________________

48. Does your **country** have an established mechanism or partnership to send patients with BCP-ALL abroad for CAR-T CD19 cells therapy?

- Yes
- No

Please write to which institutions/countries:_________________________________________

If no, select all that apply

- No, but we are looking for in country options
- No, but we are looking for international options
- No, we are not interested

49. How many children with BCP-ALL are going abroad annually for CAR-T CD19 therapy?
*(Enter 0 if none, enter 9999 if you don't have access to this data or you don't have access to this information)*
Independently/individual __________________________________________________
Using established partnerships ___________________________________________________

50. Please select all funding sources available through your **country** to support the treatment of patients with CAR-T cells therapy abroad. (select all that apply)

- Government funded
- Hospital funded
- Patient/family
- Crowd sourcing/funding
- Local Foundation/NGO
- International NGO

- Private insurance
- Other
- None

If other, please specify:_______________________________________________

51. Is your **country** legally allowed to send patients abroad from nonEMA (European Medicines Agency) approved therapy (such as an academic-based CAR-T product)?

- Yes
- No, only for EMA approved
- No
- Unknown
- Other

If other, please specify:________________________________________________

52. Please provide details about which factors (financial, regulatory, geographic, …) impact your **country's** ability to participate in a referral network for CAR-T CD19?___________

If you selected 9999 because you do not have access to data or information to answer a particular question, please provide contact information (email address) for a person with access to this data or type "Unknown" if there is no one with access to this information:____________

Please list the names of the people who contributed or collected to the data for this survey:____

Please add any additional comments related to  CAR-T CD19 cell therapy or the questionnaire below.______________________

Thank you so much for filling this questionnaire and participating in our study!

Supplementary Table 1. Reported eligibility criteria for CAR-T cell therapy in pediatric B-ALL across European countries.

Percentages indicate the proportion of surveyed countries reporting each eligibility criterion as applied in clinical practice. Abbreviations: CR, complete remission; ALL, acute lymphoblastic leukemia; allo-HSCT, allogeneic hematopoietic stem cell transplantation; TKI, tyrosine kinase inhibitor.

| Eligibility Criteria | |
|---|---|
| Failure to achieve complete remission (CR) after 2 cycles of standard chemotherapy (primary resistance for treatment) | **79%** |
| Failure to achieve CR after one cycle of reinduction therapy for relapsed ALL (secondary resistance to treatment) | **71%** |
| Second or subsequent recurrence in the bone marrow (disease recurrence) | **100%** |
| Relapse (in bone marrow) after allogeneic hematopoietic stem cell transplantation (allo-HSCT) and after an interval of at least 4 months between allo-HSCT and CAR-T administration | **100%** |
| Failure of at least two lines of tyrosine kinase inhibitor (TKI) therapy or, in the case of the EsPhALL2017 protocol, failure to achieve complete remission (CR) after consolidation. Indications also include intolerance or contraindications to TKI therapy | **58%** |
| Allo-HSCT cannot be performed (Possible causes: comorbidities, contraindications to conditioning treatment before allo-HSCT, lack of a suitable donor (e.g. only available haploidentical donor with indications only for a compatible donor according to the protocol) or previous allo-HSCT) | **54%** |
| Children above 6 months old | **29%** |
| Children less than 18 years old | **29%** |

Supplementary Table 2. Reported contraindications for CAR-T cell therapy in pediatric B-ALL across European countries

Percentages indicate the proportion of surveyed countries reporting each contraindication as applied in clinical practice. Abbreviations: ALL, acute lymphoblastic leukemia; cGvHD, chronic graft-versus-host disease.

| Contraindication | |
|---|---|
| Pregnancy or breastfeeding | 71% |
| HIV infection | 71% |
| Active hepatitis B or C | 75% |
| Actively uncontrolled systemic infection | 71% |
| Chronic graft-versus-host disease (cGvHD) stage II-IV | 63% |
| Presence of contraindications to the use of lymphodepleting chemotherapy (i.e. fludarabine and cyclophosphamide or cytarabine and etoposide) | 54% |
| Central nervous system involvement by ALL (at the time of administration of CAR-T CD19) | 21% |
| Coexistence of congenital genetic diseases with impaired bone marrow function (e.g. Fanconi anemia, Kostmann syndrome, Shwachman-Diamond syndrome) | 21% |
| Isolated extramedullary ALL recurrence | 8% |

### Supplementary Table 3.

| Country | Albania | Armenia | Austria | Belarus | Belgium | Bulgaria | Croatia | Czechia | Denmark | Estonia | Finland | France | Germany | Greece | Hungary |
|---|---|---|---|---|---|---|---|---|---|---|---|---|---|---|---|
| Income (World Bank classification (as of July 1, 2025)) | LMIC | LMIC | HIC | LMIC | HIC | HIC | HIC | HIC | HIC | HIC | HIC | HIC | HIC | HIC | HIC |
| Number of PHO centers (per country) | 1 | 1 | 5 | 3 | 7 | 3 | 1 | 7 | 4 | 2 | 5 | 27 | 55 | 6 | 7 |
| HSCT access | No | Yes | Yes | Yes | Yes | Yes | Yes | Yes | Yes | Yes | Yes | Yes | Yes | Yes | Yes |
| number of newly diagnosed pediatric B-ALL annually (per country) | 25 | 25 | 65 | 80 | 140 | 60 | 25 | 70 | 50 | 5 | 50 | 500 | 650 | NA | 70 |
| 5-year overall survival | Unknown | 75 - 100% | 75 - 100% | 75 - 100% | 75 - 100% | 75 - 100% | 75 - 100% | 75 - 100% | 75 - 100% | 75 - 100% | 75 - 100% | 75 - 100% | 75 - 100% | 75 - 100% | 75 - 100% |
| Relapse rate | Unknown | 25 - 50% | 0 - 25% | 0 - 25% | 0 - 25% | 0 - 25% | 0 - 25% | 0 - 25% | 0 - 25% | 0 - 25% | 0 - 25% | 0 - 25% | 0 - 25% | 0 - 25% | 0 - 25% |
| ALL protocol | Unknown | ALL-IC BFM | AIEOP-BFM ALL 2017 | No, we do not participate in any of these current ALL protocols | ALLTogether | ALL-IC BFM | ALL-IC BFM | AIEOP-BFM ALL 2017 | ALLTogether | ALLTogether | ALLTogether | ALLTogether | AIEOP-BFM ALL 2017 | ALL-IC BFM | ALL-IC BFM |
| availability of blinatumomab for children with BCP-ALL | Never | Almost always | Always | Always | Almost always | Always | Almost always | Always | Always | Almost always | Always | Sometimes | Always | Always | Always |
| proportion of eligible children that have access to blinatumomab as part of their BCP-ALL treatment | | 0 - 25% | 75 - 100% | 75 - 100% | 75 - 100% | 75 - 100% | 75 - 100% | 75 - 100% | 75 - 100% | 50 -75% | 75 - 100% | 75 - 100% | 75 - 100% | 75 - 100% | 75 - 100% |
| availability of allo-HSCT for children with BCP-ALL | Never | Almost always | Always | Always | Always | Almost always | Always | Always | Always | Always | Always | Always | Always | Always | Always |
| proportion of eligible children that have access to HSCT as part of their BCP-ALL treatment | | 0 - 25% | 75 - 100% | 75 - 100% | 75 - 100% | 75 - 100% | 75 - 100% | 75 - 100% | 75 - 100% | 75 - 100% | 75 - 100% | 75 - 100% | 75 - 100% | 75 - 100% | 75 - 100% |
| number of pediatric B-ALL receiving allo-HSCT annualy (per country) | NA | 3 | NA | 4 | NA | 5 | 5 | 7 | 5 | 2 | 5 | NA | 55 | 10 | 7 |
| CAR-T CD19 access | No | No | Yes | Yes | Yes | No | Yes | Yes | Yes | No | Yes | Yes | Yes | Yes | Yes |
| acceptance of patients from external centers for CAR-T cell therapy | | | Sometimes | Always | Almost always | | Almost always | Sometimes | Always | | Almost always | Sometimes | Almost always | Rarely | Sometim es |
| How many children from your country with BCP-ALL travel to another country for CAR-T CD19 therapy annually? | 2 | 3 | | | | 3 | | | | 1 | | | | | |
| Is your institution legally allowed to send patients abroad from nonEMA (European Medicines Agency) approved therapy (such as an academic-based CAR-T product)? | Unknown | Yes | Unknown | Unknown | Unknown | Unknown | Unknown | Unknown | Yes | Yes | Yes | Unknown | Yes | Unknown | Yes |

| Israel | Italy | Kazakhstan | Latvia | Lithuania | Moldova | Netherlands | Norway | Poland | Portugal | Romania | Russia | Serbia | Slovakia | Slovenia | Spain | Switzerland | Türkiye | Ukraine | United Kingdom |
|---|---|---|---|---|---|---|---|---|---|---|---|---|---|---|---|---|---|---|---|
| HIC | HIC | LMIC | HIC | HIC | LMIC | HIC | HIC | HIC | HIC | LMIC | HIC | LMIC | HIC | HIC | HIC | HIC | LMIC | LMIC | HIC |
| 6 | 41 | 2 | 1 | 1 | 1 | 1 | 4 | 15 | 3 | 35 | 80 | 4 | 3 | 1 | 40 | 9 | 50 | 26 | 22 |
| Yes | Yes | No | No | Yes | No | Yes | Yes | Yes | Yes | Yes | Yes | Yes | Yes | Yes | Yes | Yes | Yes | Yes | Yes |
| 36 | 300 | 30 | 15 | 15 | 30 | 90 | 40 | 225 | 70 | NA | 100 | 80 | 30 | 13 | 300 | 12 | NA | 150 | NA |
| 75 - 100% | 75 - 100% | Unknown | 75 - 100% | 75 - 100% | 50 -75% | 75 - 100% | 75 - 100% | 75 - 100% | 75 - 100% | 75 - 100% | 75 - 100% | 75 - 100% | 75 - 100% | 75 - 100% | 75 - 100% | 75 - 100% | 75 - 100% | 50 -75% | 75 - 100% |
| 0 - 25% | 0 - 25% | 0 - 25% | 0 - 25% | 0 - 25% | 0 - 25% | 0 - 25% | 0 - 25% | 0 - 25% | 0 - 25% | 0 - 25% | 0 - 25% | 0 - 25% | 0 - 25% | 0 - 25% | 0 - 25% | 0 - 25% | 0 - 25% | 25 - 50% | 0 - 25% |
| AIEOP-BFM ALL 2017 | AIEOP-BFM ALL 2017 | No, we do not participate in any of these current ALL protocols | ALLTogether | ALLTogether | No, we do not participate in any of these current ALL protocols | ALLTogether | ALLTogether | AIEOP-BFM ALL 2017 | ALLTogether | AIEOP-BFM ALL 2017 | No, we do not participate in any of these current ALL protocols | ALL-IC BFM | AIEOP-BFM ALL 2017 | ALL-IC BFM | ALLTogether | AIEOP-BFM ALL 2017 | ALL-IC BFM | No, we do not participate in any of these current ALL protocols | ALLTogether |
| Always | Sometimes | Sometimes | Always | Always | Never | Almost always | Always | Almost always | Always | Always | Always | Always | Almost always | Always | Sometimes | Always | Almost always | Sometimes | Always |
| 75 - 100% | 75 - 100% | 0 - 25% | 75 - 100% | 75 - 100% | | 75 - 100% | 75 - 100% | 75 - 100% | 0 - 25% | 75 - 100% | 75 - 100% | 75 - 100% | 75 - 100% | 75 - 100% | 25 - 50% | 75 - 100% | 75 - 100% | 0 - 25% | 75 - 100% |
| Always | Always | Never | Always | Always | Never | Always | Always | Always | Rarely | Always | Always | Always | Always | Always | Always | Always | Always | Always | Always |
| 75 - 100% | 75 - 100% | | 75 - 100% | 75 - 100% | | 75 - 100% | 75 - 100% | 75 - 100% | 50 -75% | 75 - 100% | 75 - 100% | 75 - 100% | 75 - 100% | 75 - 100% | 75 - 100% | 75 - 100% | 75 - 100% | 75 - 100% | 75 - 100% |
| NA | 80 | 1 | 3 | 3 | NA | 6 | 5 | 17 | 2 | 5 | 20 | 3 | 5 | 1 | NA | 25 | NA | 15 | NA |
| Yes | Yes | No | No | Yes | No | Yes | Yes | Yes | Yes | Yes | Yes | No | Yes | Yes | Yes | Yes | No | No | yes |
| Always | Sometimes | | | Always | | Sometimes | Sometimes | Always | Always | Sometimes | Never | | Rarely | Rarely | Sometimes | Sometimes | | | n/a |
| | | 0 | 0 | | 0 | | | | | | | 2 | | | | | 2 | 6 | |
| Yes | Yes | Yes | No | Yes | No | Yes | Yes | Yes | Yes | Yes | Unknown | No, only for EMA approved | No, only for EMA approved | Yes | Unknown | No | No | Yes | Unknown |
